# Efficacy of dental flossing and frequency of oral gingivitis in children and adults

**DOI:** 10.1101/2024.08.30.24312657

**Authors:** Davis Verhoeven, David Verhoeven

**Affiliations:** Ames Unified School District, Iowa State University, Ames IA; Ames, IA and Veterinary Microbiology and Preventive Medicine, Iowa State University, Ames IA

**Author notes:** Corresponding Author 1800 Christensen Dr, Iowa State University, College of Veterinary Medicine, VMPM, Ames, IA 50011.

## Abstract

Tooth brushing and flossing are usually both hallmarks of a good oral hygiene routine to prevent decay, gingivitis, and periodontitis. While brushing removes much of the oral bacteria from the front and backs of the teeth, flossing is believed to be necessary to remove bacteria between the teeth. However, the effectiveness of self-flossing has not been established very well. Flossing effectiveness was evaluated two ways in this study: adults and 12 year old children were instructed on how to floss and bacterial colonies were determined before or after 7 days or pediatric and family dentists were blinded to patient surveys that asked about flossing frequency and evaluated the patient for gingivitis. We found a significant number of children did not floss at all despite brushing daily. However, flossing had no effect on the number of bacteria in their mouths nor did flossing have any correlation with reducing gingivitis development. Taken together, self-flossing did not appear to be an effective strategy for reduction of gingivitis in children or adults but could stem from improper technique or simply a lack of doing it.

## Introduction

A tenet of maintaining oral health is daily tooth brushing, flossing, and at least an annual, if not biannual, preventative cleaning by a dental professional. Flossing between the teeth is believed to manually remove bacteria that inhabits that area and is difficult to remove by brushing alone. However, the data supporting the effectiveness of self-oral flossing is not well supported although there are studies suggeting its important to overall oral health (1).

Furthermore, the effectiveness of flossing in children has not been well studied as an additive step in brushing to prevent/reduce oral gingivitis although the technique used might be imporant (2). Gingivitis is a serious inflammatory reaction to oral flora marked by red inflamed gum lines with potential to leading to bleeding and gum line recession (3). In cases of untreated gingivitis, periodontitis can develop which leads to severe gum disease with potential loss of bone that supports teeth. Thus, maintaining good oral hygiene is paramount to preventing tooth loss and oral decay. Again, whether self-flossing contributes to prevention of the initial catalyst of periodontitis is just not well known.

The oral microbiota is composed of hundreds to thousands of bacterial species that colonize the teeth and gum lines usually as biofilms and serve to exclude more pathogenic oral pathogens from taking root (4, 5). The largest groups of commensal bacteria belong to the species *Streptococcus, Actinomyces, Veillonella, Fusobacterium, Porphromonas, Prevotella, Treponema, Nisseria, Haemophilis, Eubacteria, Lactobacterium, Capnocytophaga, Eikenella, Leptotrichia, Peptostreptococcus, Staphylococcus*, and *Propionibacterium*. Generally, these species of bacteria live in harmony with their host and do not often invoke immune reaction at the gum line that lead to gingivitis. In contrast, colonization on the teeth and in the gums of species *A. actinomycetemcomitans, P. gingivalis, T. forsythensis, Porphyromonas* spp., *Campylobacter* spp., *or Treponema denticola* will almost always lead to gingivitis as these bacteria are quite invasive and immunogenic. However, more pathogenic bacterial are not alone in causing disease as chronic gingivitis can also result from changes in biofilms comprised mainly of gram-positive facultative anaerobes (*Streptococcus anginosus* and *A. naeslundii*) as well. Thus, removal of pathogenic oral pathogens that attempt to gain access to area around the gum line through flossing as a supplement to brushing is thought to contribute to a reduction in gingivitis. Maintaining flossing should also, in theory, reduce bacterial overgrowth of the commensal bacteria at the tops biofilms to again reduce the incidence of gingivitis from the gram-positive bacteria.

Here, we examined the efficacy of self-flossing at reducing the incidence of oral gingivitis. We found that both adults and children had no change in the number or type of bacteria colonizing their teeth before or after flossing. Furthermore, no correlation was found between flossing and the development of gingivitis as determined by dental professionals. Thus, this study demonstrates that self-flossing may not be effective at preventing gingivitis and further stresses the importance of professional cleanings by trained dental professionals. Alternatively, people may not be flossing correctly/adequately to remove bacteria between the teeth and better training by dentists/hygienists may be needed.

## Methods and Materials

### Subjects

Children (aged 11-13 years old, n=15) and adults (aged 25-50 years old, n=15) without compounding dental diseases were recruited for the flossing and bacterial burden portion of the study. Additional children (7-13 years old) and adults (20-55 years old) were recruited to the study from local (Ames IA) pediatric dental or family dental practices. All procedures were approved by Ames Unified IRB. Sample identification were blinded to the researchers for analysis. Dentists were blinded to whether patients flossed or did floss as self-identified on their surveys. The communities that this study draws from have about 0.35mg/liter of fluoride naturally in the drinking water.

### Testing Procedure

Sterile floss marked at two points was supplied to subjects who proceeded to floss the top and bottom teeth. Subjects were instructed to keep fingers outside the marks upon the floss as these points were cut to create a floss segment, containing organisms with origins from the mouth (Figure 1). The floss segments were deposited into a 1.5ml tube containing 500µl of steile phosphate buffered saline (PBS) and collected immediately after sampling and held in a four degree C refrigerator for < 3 hours before processing. Subjects were instructed to floss for one week prior to collection of oral bacteria (post-flossing sampling). After this period, the testing procedure listed above was repeated.

**Figure 1.**
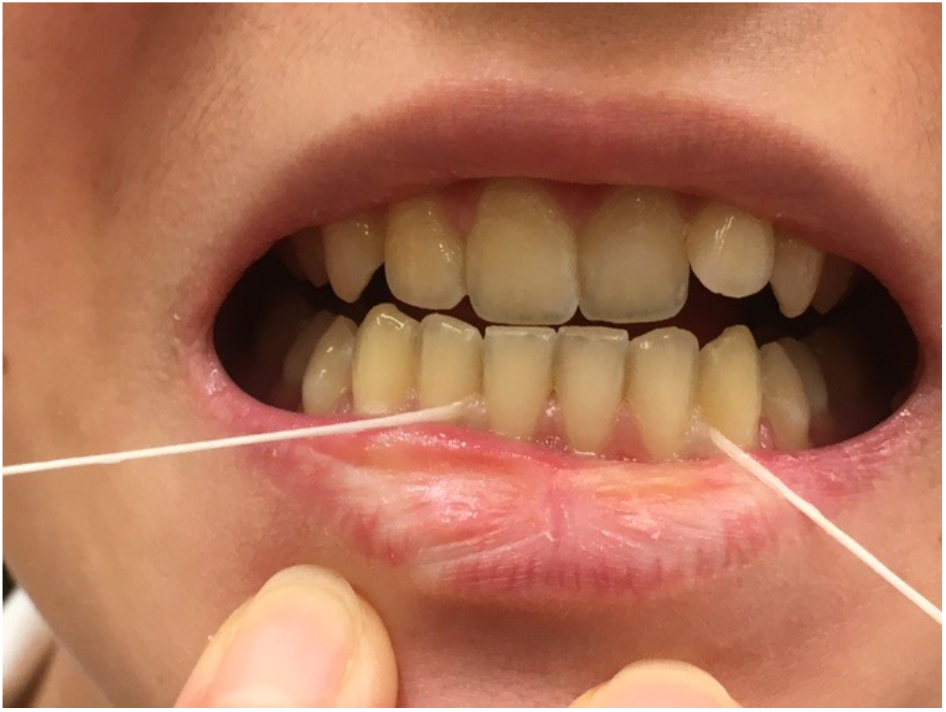
Example of our technique. (Left) (Right) Lines show areas of teeth sampled for bacteria pre or after 1 week of flossing. Flossing from all four surfaces were pooled for bacterial counts.

### Bacterial culturing

The PBS solution containing floss from subjects was subjected to vortexing, for a minimum of 30 seconds. Serial dilutions in PBS were plated and spread on Tryptic Soy petri dishes with 5% sheep blood. This bacteria was cultured aerobically for 24 hours in 37C. Bacteria was also plated from the same source was cultured for 240 hours anaerobically at 37C using anaerobic sleeves (BD). After culturing, the amount of bacteria was recorded in colony forming units (CFUs) using dilution plating for both culturing conditions irrespective of bacterial type.

### Bacterial Identification

To identify the types of bacteria growing in between teeth, PCR on spun samples were used. To identify the bacteria, universal r16s primers were used to amplify and bacteria and subjected to Sanger sequencing from bacterial colonies. To test for the specific species of bacteria associated greatest with Gingivitis, multiplex PCR was also used to identify A. *actinomycetemcomitans*, P. *gingivalis*, and T. *forsythensis*.

### Statistical analysis

Statistics were calculated using Prism Software Utilizing a 2×2 contingency table or a Chi-Test for correlation or paired T-tests. A p-value of 0.05 or less was considered significant.

## Results

### No correlation between self-flossing and diagnosis of gingivitis

To examine the level of gingivitis in children and adults at their annual or biannual dental check-up, patient self-identified as to the frequency of self-flossing on a survey and were assigned a number. The patients’ answers were blinded to the dentist who indicated whether the patient exhibited gingivitis or not during their routine exam of their oral health. After results were unblinded, there were no correlations between whether a child or adult had gingivitis and whether they flossed (Table 1). The subjects were then further broken into groups based on frequency of flossing and survey results reexamined for presence of gingivitis. As shown in Table 2, there again was no correlation between gingivitis and the frequency of flossing with patients equal likely to have gingivitis whether they were frequent, infrequent, or never flossed.

**Table 1.** Survey of flossing habits and gingivitis fails to establish a correlation between flossing and prevention of gingivitis. Dentists were blinded to their patients flossing habits. Each dentist, mixed practice and pediatric dentist, assessed whether gingivitis was present in their patients or not.

All individuals
| Flossing with no gingivitis | Flossing but has gingivitis |
| --- | --- |
| 28 | 24 |
| No flossing with no gingivitis | No flossing but has gingivitis |
| 13 | 5 |

p-value 0.26 Not significant

Children (<15 years old)
| Flossing with no gingivitis | Flossing but has gingivitis |
| --- | --- |
| 22 | 12 |
| No flossing with no gingivitis | No flossing but has gingivitis |
| 11 | 5 |

p-value 1.0 Not significant

| Flossing with no gingivitis | Flossing but has gingivitis |
| --- | --- |
| 6 | 12 |
| No flossing with no gingivitis | No flossing but has gingivitis |
| 2 | 0 |

p-value 0.17 Not significant

**Table 2.**
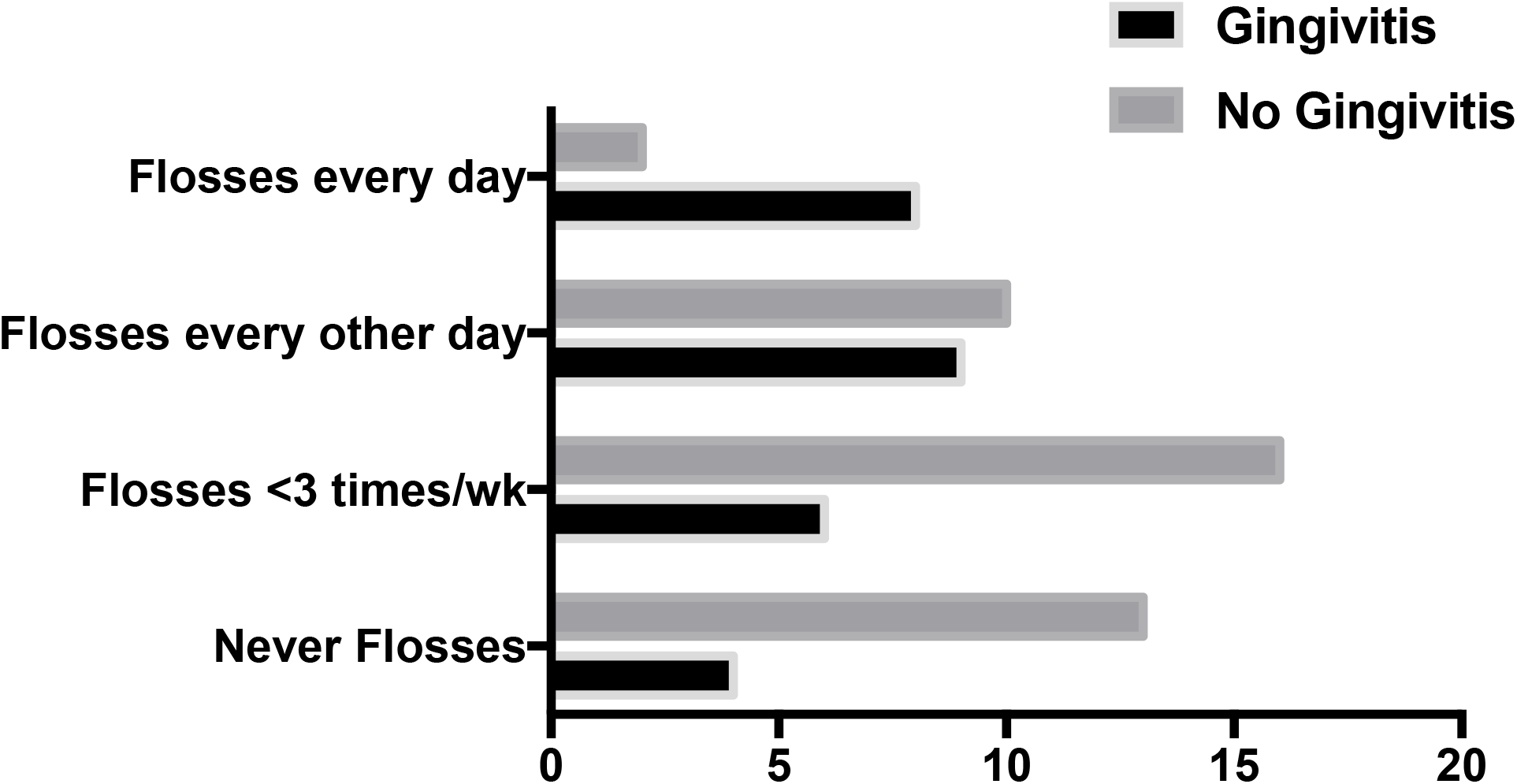
Frequency of flossing also shows no correlation between more flossing or less and prevention of gingivitis. Patients were also assessed as to frequency of flossing prior to dental evaluation through an intake survey form given to each one.

### Self-flossing is not effective at reducing bacterial burdens in adults or children

Since there did not appear to be a correlation between flossing and the level of gingivitis, we next surveyed for whether children or adults were reducing the number of colonizing bacteria between their teeth before or after flossing. Subjects who identified as never flossing were sampled for bacteria between their teeth and then instructed on how to floss for a 2 week period. They were then retested after this period with bacteria plated and counted pre and post flossing. There was no significant change in the number of bacteria isolated after the subjects had been flossing after having not flossed prior (Figure 2). In two additional adult patients that were regular flossers and had their teeth sampled prior to stopping flossing for two weeks, no difference in their bacterial numbers was detected (data not shown).

**Figure 2.**
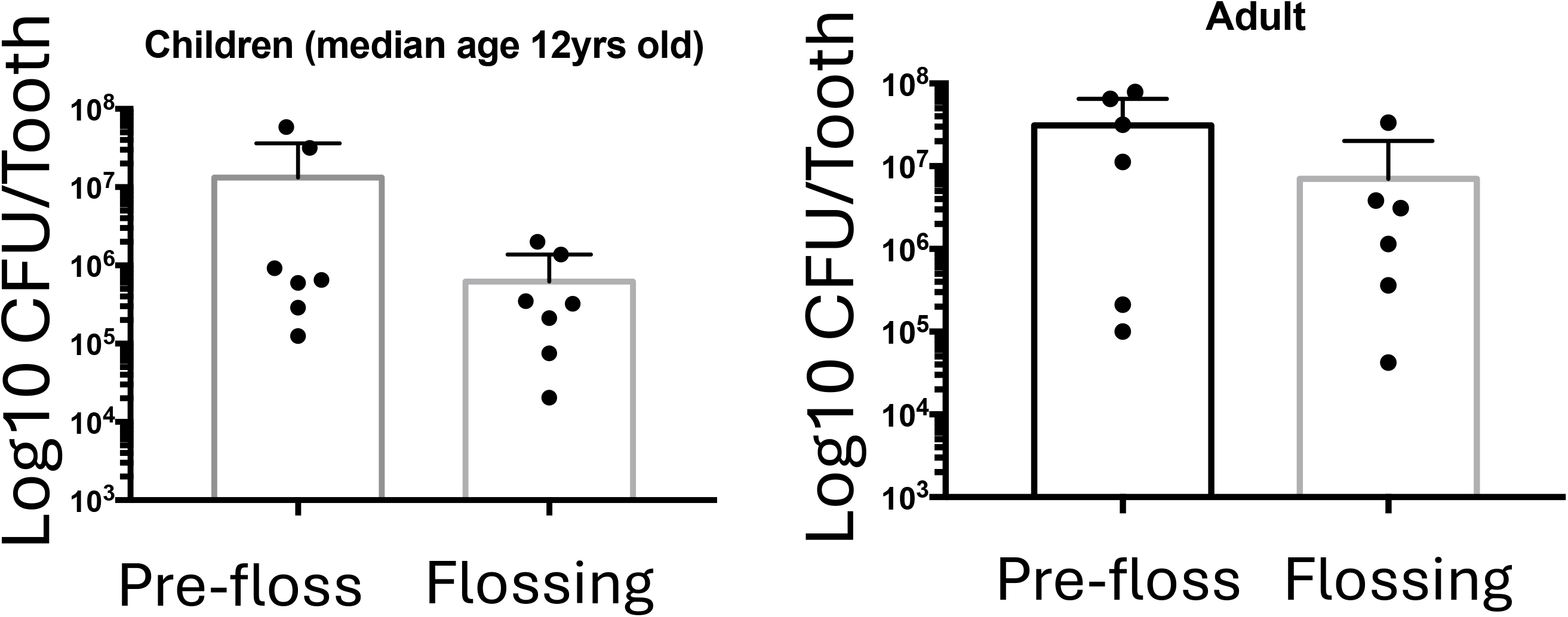
The number of bacterial colonies over all individuals or as a comparison between children and adults. This graph shows the different number of colonies present for each subject on each plate with median. Dots represent CFU (colony forming unit) counts per tooth flossed. CFUs for children are in the upper graph and adults are in the lower graph. p-values calculated using paired T-test.

### Self-flossing does not change the type of bacteria between the teeth

Since flossing did not change the number of bacteria, we next examined whether flossing changed the type of bacteria colonizing between the teeth. However, flossing, like the number of total bacterial, did not appear to change the type of bacteria isolated by methodologies. Specifically, we found a majority of the bacteria species did not change pre or during flossing as determined with 16s sequence showing no differences in *Strep* species, or other bactieral species detected (i.e. *Fusobacterium, Leptotrichia, Campyobacter*, etc).

### Flossing fails to prevent oral pathogens from colonizing between the teeth

Among the common gingivitis-causing pathogens, *P. gingivalis* and *T. forsythensis* were detected in some patients’ pre and post flossing. We did not detect some of these pathogens prior to flossing but did after. Overall these data suggest that flossing does not affect the colonization of known oral pathogens either.

## Discussion

Flossing has long been suggested by dental professionals as a way to remove plaque and bacterial buildup between the teeth. However, the effectiveness of self-flossing has never been fully proven that we are aware of. In fact, a search of google shows many dental websites stating the need for flossing but little in actual studies to support it. Here, we sought to determine whether self-flossing in adults and children could achieve a reduction in bacteria between the teeth and whether routine flossing correlates with a reduced incidence of gingivitis. This study suggests that self-flossing may not be that effective as a supplement to brushing in adults or children.

The study does a few limitations. First, obviously a larger sample size would certainly help bolster our findings although this study was adequately powered. Second, we did not have the dentist grade the scale of gingivitis but rather relied on a diagnosis of gingivitis present or absent. Thirdly, a more robust removal of bacteria between the teeth might help identify any changes in oral biofilms than the methods used in this study. We also did not do deep sequencing of oral biofilms in between the teeth. However, we believe that flossing would unlikely remove much of the biofilms. Finally, we did not test the effectiveness of flossing with toothpaste directly in the mouth and it is feasible that flossing could push paste in between the teeth better and reduce bacteria that way.

The effectiveness of self-flossing may come down to poor technique. We were surprised at how many of our younger subjects never flossed. In the few subjects where bacteria strains like *T. forsythensis*, that may come down to sampling technique. Likely with the start of flossing, commensals were now removed that allowed more traditional bacteria to grow. While we only concentrated on correlations between oral self flossing and gingivitis, flossing could still help prevent other tooth decay or plaque buildups. There are hints in prior studies that it also may not be that effective for plaque in adults (6). Flossing with other oral aides may be prove more effective (7) or better techniques (8). All together, these data strongly advocate for professional cleaning to maintain oral health.

## Data Availability

All data produced in the present study are available upon reasonable request to the authors

## Acknowledgements

We thank our participating dentists, their hygienist staff, and patients for participating in this study.

